# Epigenetic Age and Lung Cancer Risk in the CLUE II Prospective Cohort Study

**DOI:** 10.1101/2022.05.20.22275270

**Authors:** Dominique S. Michaud, Mei Chung, Naisi Zhao, Devin C. Koestler, Jiayun Lu, Elizabeth A. Platz, Karl T. Kelsey

## Abstract

**Background:** Epigenetic age, a robust marker of biological aging, has been associated with obesity, low-grade inflammation and metabolic diseases. However, few studies have examined associations between different epigenetic age measures and risk of lung cancer, despite great interest in finding biomarkers to assist in risk stratification for lung cancer screening.

**Methods:** A nested case-control study of lung cancer from the CLUE II cohort study was conducted using incidence density sampling with 1:1 matching of controls to lung cancer cases (n=208 matched pairs). Prediagnostic blood samples were collected in 1989 (CLUE II study baseline) and stored at −70°C. DNA was extracted from buffy coat and DNA methylation levels were measured using Illumina MethylationEPIC BeadChip Arrays. Three epigenetic age acceleration (i.e., biological age is greater than chronological age) measurements (Horvath, Hannum and PhenoAge) were examined in relation to lung cancer risk using conditional logistic regression.

**Results:** We did not observe associations between the three epigenetic age acceleration measurements and risk of lung cancer overall; however, inverse associations for the two Hannum age acceleration measures (intrinsic and extrinsic) were observed in men and among younger participants, but not in women or older participants. We did not observe effect modification by time from blood draw to diagnosis.

**Conclusion:** Findings from this study do not support a positive association between three different biological age acceleration measures and risk of lung cancer. Additional studies are needed to address whether epigenetic age is associated with lung cancer in never smokers.

## Introduction

Lung cancer remains the leading cause of cancer deaths in the US and worldwide [1]. Substantial effort has been devoted to identifying heritable genomic markers that could aid in classification of high-risk individuals for screening purposes [2, 3]. While results from these studies are promising, predictive modeling using genomic markers (in addition to age and smoking) are currently not sufficiently discriminatory or calibrated to be useful in clinical settings for risk prediction. Identifying high risk groups could improve efficiency in lung cancer screening (with LDCT) and reduce racial inequalities associated with the current recommendations for screening based on smoking history [4]. Thus, there is an urgent need to identify biomarkers that can reflect biological processes in lung cancer development and that could, eventually, be incorporated into models for risk stratification.

Variation in DNA methylation levels in peripheral blood leukocytes reflect genetic imprinting, environmental exposures, and the lineage differentiation that gives rise to immune cell subtypes [5]. Recent studies using epigenetic markers in blood have identified differentially methylated regions in smokers [6, 7]; DNA methylation levels in these regions remained strongly associated with lung cancer mortality after adjusting for smoking history [6]. Epigenetic aging measures or “clocks” have also been developed to reflect biological age in tissue and blood [8]. These epigenetic clocks are highly correlated with chronological age, but can deviate from chronological age, reflecting changes in immunity and cellular senescence, which are closely aligned with health and disease. Epigenetic age acceleration occurs when the predicted biological age is greater than chronological age. Recent studies have linked epigenetic age acceleration to a range of disease outcomes, including all-cause mortality [9, 10], cardiovascular disease (CVD) incidence [11], CHD mortality [12], cancer incidence [13] and cancer mortality [12]. Epigenetic age acceleration estimated using the Horvath and Hannum clocks, known as “first generation clocks”, is highly heritable (∼0.4 [9]) and has been associated with CVD and cancer risk factors, including obesity, low-grade inflammation [14], and metabolic syndrome [15].

To date, studies evaluating epigenetic age acceleration and lung cancer risk have been inconsistent. The first nested case-control study conducted in the Women’s Health Initiative (WHI) observed a strong positive association [16], while a larger nested case-control study (Melbourne Collaborative Consortium Study; MCCS) reported no associations for the Horvath and Hannum clocks [17]. In the WHI study, the positive associations were stronger among women developing lung cancer at 70 or more years and among current smokers. In the MCCS [17], men and women were combined, and no stratified analyses were conducted by sex, to inform whether the association was restricted to women.

It is important to examine whether epigenetic age is associated with lung cancer risk across multiple prospective studies to determine its utility as a potential biomarker to be considered for risk stratification in the selection of high-risk individuals for lung cancer screening. Thus, for this analysis, we conducted a nested case-control analysis of 208 lung cancer cases and 208 matched controls with archived pre-diagnostic blood samples (from 1989). The case-control study is nested in the CLUE II cohort study, a predominantly White cohort of men and women, based in Maryland, USA.

## Methods

### Study Population

Individuals included in this analysis were selected from participants in the CLUE II study, a prospective cohort study initiated in Washington County, Maryland, in 1989 [18, 19]. The CLUE II study was an outgrowth of a previous study (CLUE I) that had been conducted in the same region in 1974. Some of the participants in CLUE II had been participants in CLUE I (about a third), but this was not a requirement for recruitment into the overall CLUE II cohort. At the baseline visit (1989 for CLUE II), brief medical histories, blood pressure readings, and blood samples were collected on 32,894 participants (25,076 of which were residents of Washington County). Mobile office trailers were used to recruit participants and to collect blood samples. Blood was drawn into 20 ml heparinized Vacutainers (Becton-Dickinson, Rutherford, NJ), kept at 4°C until the plasma was separated, usually within 2–6 h, and divided into aliquots of plasma, buffy coat, and red blood cells. All samples are stored at −70°C. Comparisons with published figures from the 1990 Census indicated that approximately 30 percent of adult residents had participated: 98.3% were White, reflecting the population of this county, and 59% were female, with the better-educated and the age group 45 to 70 years having higher participation rates. Self-reported attained education, weight and height, cigarette smoking status, number of cigarettes smoked per day, and cigar/pipe smoking status were recorded for each participant at baseline.

The Institutional Review Board at the Johns Hopkins Bloomberg School of Public Health and the Tufts University Health Sciences Campus Institutional Review Board approved this study.

### Lung cancer cases and matched controls

Incident lung cancer cases were ascertained from linkage to the Washington Co. Cancer Registry (1989-January 2018) and the Maryland Cancer Registry (1992-January 2018). The Maryland Cancer Registry is certified by the North American Association of Central Cancer Registries as being more than 95% complete. Compared with the Maryland Cancer Registry, the Washington County Cancer Registry captured 98% of the lung cancer cases diagnosed in Washington County residents in 1998. Cancer deaths were identified from state vital statistics, next of kin, and obituaries and confirmed on death certificates; underlying cause of death was obtained from the death certificates. Between 1989 and January 2018, a total of 241 eligible incident first primary lung cancer cases were ascertained from CLUE II participants with blood samples, and who had also previously participated in CLUE I (a requirement based on a shared study population). All 241 lung cancer cases (ICD 9 162 and ICD10 C34) were confirmed by pathology report.

Controls were selected from among CLUE II participants who had also participated in CLUE I. Matching was conducted using incidence density sampling such that a control had to be alive and free of cancer at the time the matched case was diagnosed with lung cancer. One control was matched to each case on the following factors: age (±3 year), sex, race, cigarette smoking status and intensity, cigar/pipe smoking status, and date of blood draw (±4 months). Controls who later became cases were also included as cases with their new matched controls.

### DNA methylation measurements

Extracted DNA was bisulfite-treated using the EZ DNA Methylation Kit (Zymo), and DNA methylation was measured with the 850K Illumina Infinium MethylationEPIC BeadChip Arrays (Illumina, Inc, CA, USA). All samples and all array measurements were performed blinded to case-control status. Details on DNA methylation measurements, data preprocessing processing and quality control assessment/screening have been published [20]. Due to lack of remaining DNA, 8 of the 241 incident cases were removed from the dataset before matching.

### Estimation of peripheral blood leukocyte composition

Peripheral blood leukocyte subtypes proportions were estimated using a newly expanded reference-based deconvolution library EPIC IDOL-Ext [21]. This library used the IDOL methodology [22] to optimize the currently available six-cell reference library [23] to deconvolute the proportions of 12 leukocyte subtypes in peripheral blood (neutrophils, eosinophils, basophils, monocytes, naïve and memory B cells, naïve and memory CD4+ and CD8+ cells, natural killer, and T regulatory cells).

### Data processing

All methylation data preprocessing and normalization steps were performed using the Bioconductor packages. The raw IDAT files from methylation array were processed using the minfi Bioconductor package [24, 25]. Within-array correction for background fluorescence and dye-biases were performed using the Noob methodology via the function “preprocessNoob” in the minfi Bioconductor package [26]. The QCinfo function in ENmix Bioconductor [27] package was then used to identify and remove poor quality samples and probes. Samples were excluded if: 1) more than 5% of probes had quality issues as addressed using the detection p-value, 2) the bisulfite conversion intensity was lower than 3 standard deviations from the mean, or 3) the mean average intensity and/or the mean average beta values were more than 3 times IQR from the upper quartile or less than 3 time IQR from the lower quartile of the average intensity values or beta value across the samples. In addition, we excluded probes that had detection p-values exceeding 1×10^−6^ (compared to the negative background probes) in more than 5% of the samples. After sample- and probe-level quality control, we corrected the type II probe bias to make the methylation distribution of type II feature comparable to the distribution of type I feature using the beta mixture quantile dilation intra-sample normalization method [28], implemented using “BMIQ” function in the wateRmelon Bioconductor package [29]. Principal components analysis (PCA) was performed on the BMIQ-adjusted values and the top K principal components (K determined using a previously described random matrix theory approach [30]) to detect whether the microarray dataset had the batch effect. Then *ctrlsva* function in ENmix Bioconductor packages [31] was used to estimate the surrogate variables of batch effects [32]. The estimated surrogate variables were used in downstream analyses to adjust for batch effects and other unwanted technical sources of variation.

### Pack-years methylation score

A pack-years methylation score was calculated to represent pack-years smoked-associated methylation alterations [33]. The pack-years methylation score was first developed to predict smoking pack-years using smoking ‘signatures’ reported by large-scale epigenome-wide association meta-analyses [33]. The score correlates with gene expression changes affected by smoking and can be utilized in lieu of self-reported smoking data.

### Estimation of epigenetic age

Three DNAm clocks (Hannum [34], Horvath [35], and PhenoAge [36]) were used to estimate subjects’ DNAm age (using ENMix Biocondontor package: https://rdrr.io/bioc/ENmix/man/methyAge.html). For each of the three DNAm clocks, DNAm age acceleration (AA) was defined by regressing DNAm age on chronologic age and calculating the difference between the observed chronological age and the fitted DNAm age (i.e., the residual). Additionally, intrinsic epigenetic age acceleration (IEAA) metrics were calculated using the residuals from the linear regression fit to DNAm age on chronologic age, adjusted for estimated blood cell composition [37, 38] (for comparability with prior studies, we did not update the reference library for the IEAA measurements). Three subjects with an absolute value of the age acceleration estimate greater than 3 standard deviations (SDs) from the mean were excluded.

### Statistical analyses

Given the 1:1 case-control matching present in our study, conditional logistic regression models were used to examine the association between epigenetic age acceleration and lung cancer risk. As age, sex, and smoking status (never, former, current), smoking intensity (cigarettes/day) and cigar/pipe smoking were matching factors, these were implicitly adjusted for when using conditional regression. In the regression model, we additionally adjusted for BMI as a continuous variable, batch effect (for methylation arrays), and a previously described methylation-predicted variable to capture pack-years smoked [39]. We conducted stratified analyses by sex, median age, lung cancer histology (non-small cell lung cancer [NSCLC], small cell lung cancer [SCLC]), and length of time between blood draw and diagnosis (<=10, >10 years) to evaluate potential effect modification. We did not adjust for methylation-derived cell proportions given that the intrinsic epigenetic age (IEAA) measures already account for immune cells. Pearson’s correlation was used to examine correlation between epigenetic age acceleration and methylation predicted immune cell proportions. All statistical analyses were performed in R (version 3.5.1).

## Results

The final analysis consisted of 208 cases and matched 208 controls. As a result of matching, lung cancer cases and controls had similar age (mean age: 55.9 years among controls, 58.3 years among cases), sex distribution (54.3% females in cases and controls), smoking status (51% current smokers in cases and controls; 39% former smokers in cases and controls), smoking intensity (25 cigarettes per day in current smokers among cases; 24 cigarettes/day in current smokers among controls) and cigar or pipe smoking (15% ever in cases and controls). Only 3 cases, and no controls, were non-White individuals. Cases and controls were also similar with respect to BMI (mean, in kg/m^2^, BMI 26.0 cases, 26.2 controls). Most lung cancer cases were NSCLC (74%). Cases were diagnosed a mean of 14 years post-blood donation (median 14 years; range >0-29 years; all cases were incident cases).

In this population, age acceleration with all 3 measures was more common among cases with a shorter time between blood draw and cancer diagnosis (mean 13 vs. 15 for years) and among men (Table 1). Other characteristics, including smoking and BMI were very similar for acceleration and deceleration of epigenetic age in all 3 measures.

**Table 1.**
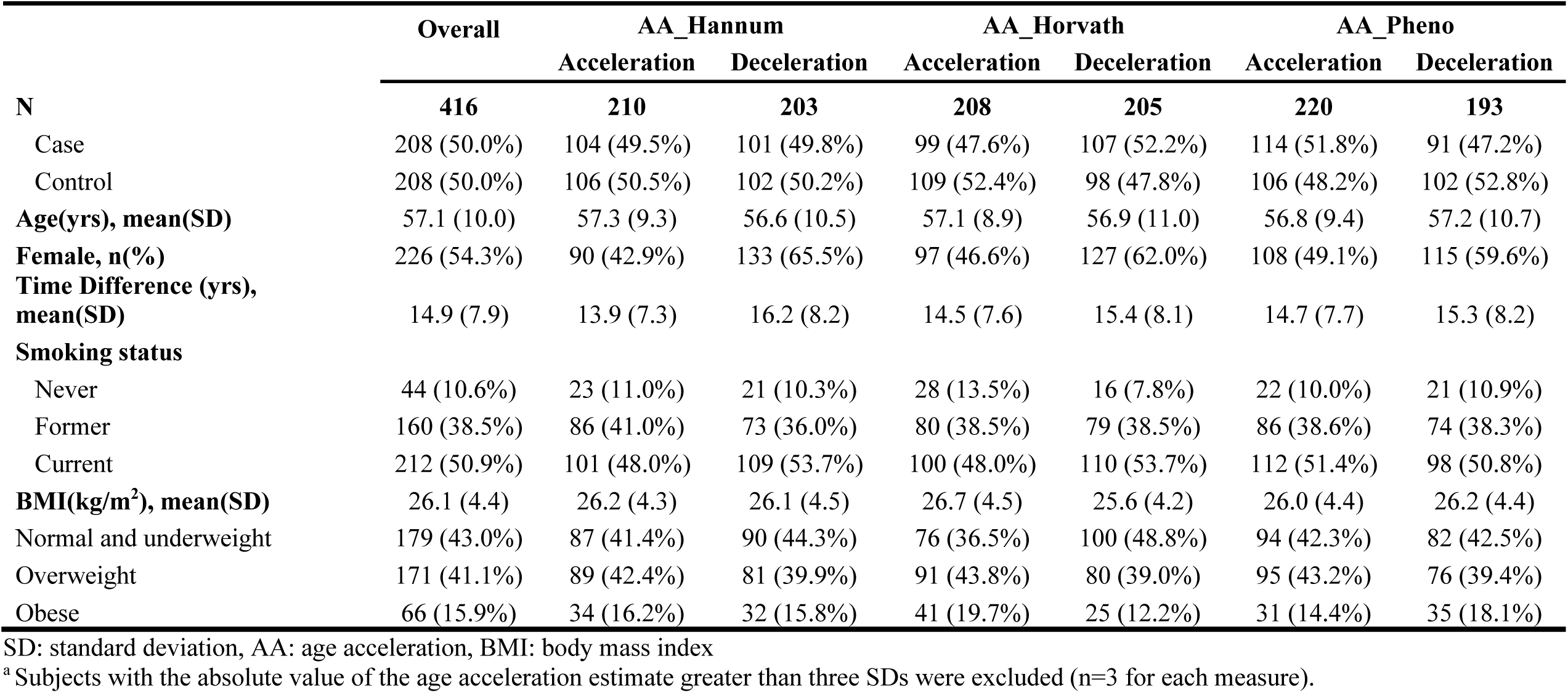
Baseline characteristics for the CLUE II population, by age acceleration vs. deceleration ^a^.

|  | Overall | AA_Hannum |  | AA_Horvath |  | AA_Pheno |  |
| --- | --- | --- | --- | --- | --- | --- | --- |
|  |  | Acceleration | Deceleration | Acceleration | Deceleration | Acceleration | Deceleration |
| **N** | **416** | **210** | **203** | **208** | **205** | **220** | **193** |
| Case | 208 (50.0%) | 104 (49.5%) | 101 (49.8%) | 99 (47.6%) | 107 (52.2%) | 114 (51.8%) | 91 (47.2%) |
| Control | 208 (50.0%) | 106 (50.5%) | 102 (50.2%) | 109 (52.4%) | 98 (47.8%) | 106 (48.2%) | 102 (52.8%) |
| **Age(yrs), mean(SD)** | 57.1 (10.0) | 57.3 (9.3) | 56.6 (10.5) | 57.1 (8.9) | 56.9 (11.0) | 56.8 (9.4) | 57.2 (10.7) |
| **Female, n(%)** | 226 (54.3%) | 90 (42.9%) | 133 (65.5%) | 97 (46.6%) | 127 (62.0%) | 108 (49.1%) | 115 (59.6%) |
| **Time Difference (yrs), mean(SD)** | 14.9 (7.9) | 13.9 (7.3) | 16.2 (8.2) | 14.5 (7.6) | 15.4 (8.1) | 14.7 (7.7) | 15.3 (8.2) |
| **Smoking status** |  |  |  |  |  |  |  |
| Never | 44 (10.6%) | 23 (11.0%) | 21 (10.3%) | 28 (13.5%) | 16 (7.8%) | 22 (10.0%) | 21 (10.9%) |
| Former | 160 (38.5%) | 86 (41.0%) | 73 (36.0%) | 80 (38.5%) | 79 (38.5%) | 86 (38.6%) | 74 (38.3%) |
| Current | 212 (50.9%) | 101 (48.0%) | 109 (53.7%) | 100 (48.0%) | 110 (53.7%) | 112 (51.4%) | 98 (50.8%) |
| **BMI(kg/m<sup>2</sup>), mean(SD)** | 26.1 (4.4) | 26.2 (4.3) | 26.1 (4.5) | 26.7 (4.5) | 25.6 (4.2) | 26.0 (4.4) | 26.2 (4.4) |
| Normal and underweight | 179 (43.0%) | 87 (41.4%) | 90 (44.3%) | 76 (36.5%) | 100 (48.8%) | 94 (42.3%) | 82 (42.5%) |
| Overweight | 171 (41.1%) | 89 (42.4%) | 81 (39.9%) | 91 (43.8%) | 80 (39.0%) | 95 (43.2%) | 76 (39.4%) |
| Obese | 66 (15.9%) | 34 (16.2%) | 32 (15.8%) | 41 (19.7%) | 25 (12.2%) | 31 (14.4%) | 35 (18.1%) |

SD: standard deviation, AA: age acceleration, BMI: body mass index
<sup>a</sup> Subjects with the absolute value of the age acceleration estimate greater than three SDs were excluded (n=3 for each measure).

Overall, we did not observe any associations between the 3 epigenetic age acceleration measures and lung cancer risk using both continuous and categorical variables for age acceleration (Table 2). Associations were similar when stratified by follow-up time (Table 3).

**Table 2.** Odds ratios and 95% confidence intervals for the association between epigenetic age acceleration (using 3 different measures) and the risk of lung cancer in the CLUE II study.

| Age acceleration measure | OR (95% CI)* | p-value* |
| --- | --- | --- |
| AA_Hannum |  |  |
| Q1 | 1 (ref.) |  |
| Q2 | 0.96 (0.55, 1.66) | 0.88 |
| Q3 | 0.74 (0.42, 1.30) | 0.29 |
| Q4 | 0.74 (0.42, 1.31) | 0.31 |
| Continuous (per 1 SD) | 0.82 (0.64, 1.05) | 0.12 |
| AA_Horvath |  |  |
| Q1 | 1 (ref.) |  |
| Q2 | 0.80 (0.46, 1.41) | 0.45 |
| Q3 | 0.65 (0.37, 1.16) | 0.15 |
| Q4 | 0.76 (0.43, 1.33) | 0.33 |
| Continuous (per 1 SD) | 0.85 (0.68, 1.07) | 0.17 |
| AA_Pheno |  |  |
| Q1 | 1 (ref.) |  |
| Q2 | 0.61 (0.33, 1.11) | 0.10 |
| Q3 | 0.79 (0.44, 1.40) | 0.42 |
| Q4 | 0.89 (0.51, 1.56) | 0.69 |
| Continuous (per 1 SD) | 0.92 (0.76, 1.10) | 0.36 |
| IEAA_Hannum |  |  |
| Q1 | 1 (ref.) |  |
| Q2 | 0.74 (0.42, 1.31) | 0.30 |
| Q3 | 0.62 (0.35, 1.10) | 0.10 |
| Q4 | 0.70 (0.40, 1.24) | 0.22 |
| Continuous (per 1 SD) | 0.87 (0.67, 1.14) | 0.33 |
| IEAA_Horvath |  |  |
| Q1 | 1 (ref.) |  |
| Q2 | 0.52 (0.28, 0.94) | 0.031 |
| Q3 | 0.84 (0.48, 1.47) | 0.54 |
| Q4 | 0.82 (0.47, 1.42) | 0.47 |
| Continuous (per 1 SD) | 0.93 (0.73, 1.19) | 0.58 |
| IEAA_Pheno |  |  |
| Q1 | 1 (ref.) |  |
| Q2 | 0.75 (0.42, 1.35) | 0.33 |
| Q3 | 0.92 (0.52, 1.65) | 0.79 |
| Q4 | 0.87 (0.49, 1.55) | 0.64 |
| Continuous (per 1 SD) | 0.90 (0.74, 1.10) | 0.30 |

IEAA: Intrinsic epigenetic age acceleration
\*Conditional logistic regression models, adjusting for batch effects, BMI, and smoking predicted packyears (using DNA methylation).

**Table 3.**
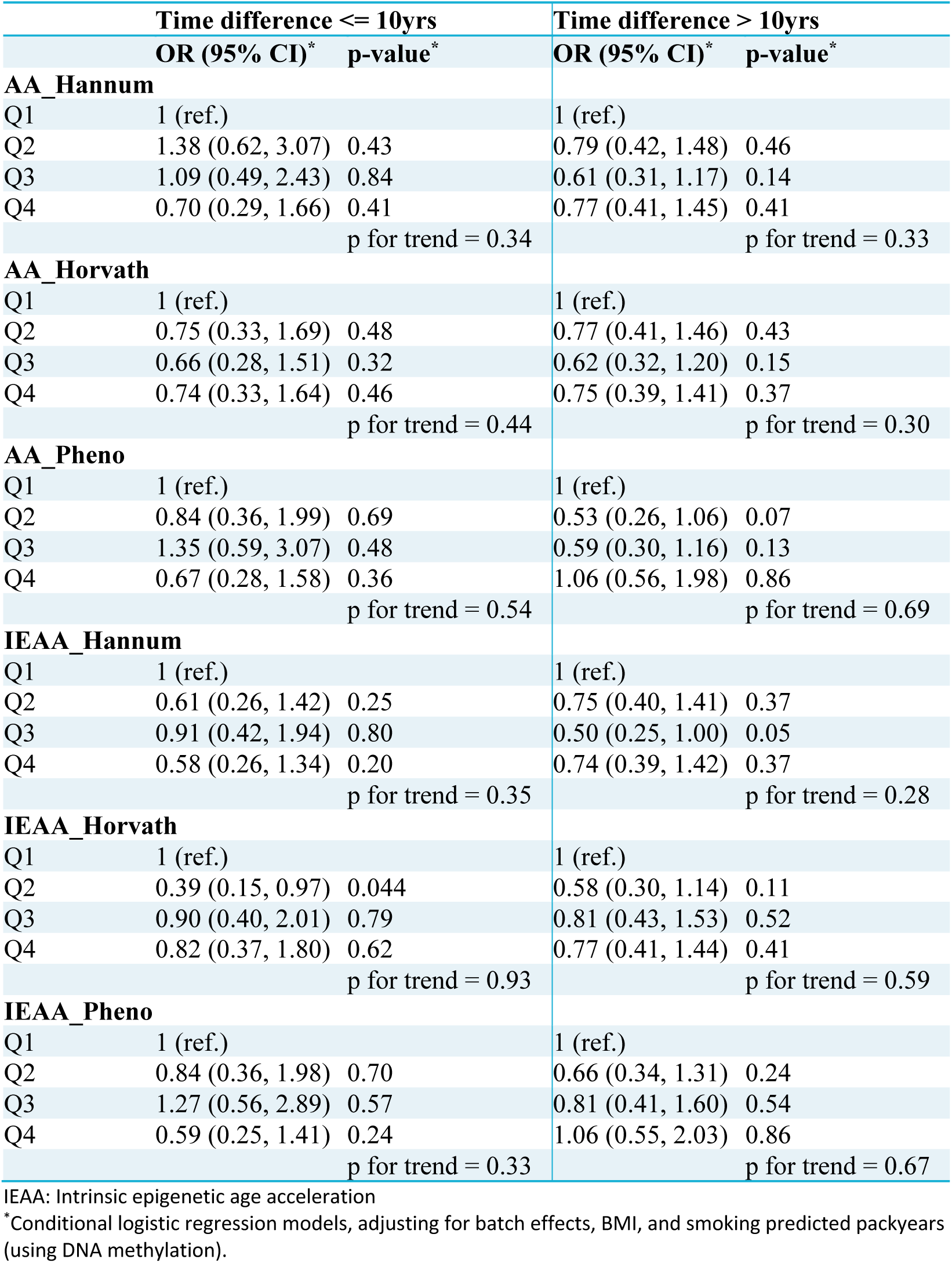
Odds ratios and 95% confidence intervals for the association between epigenetic age acceleration (using 3 different measures) and the risk of lung cancer in the CLUE II study, stratified by time between blood draw and cancer diagnosis (in cases; matched date in controls).

Given that in a prior study positive associations were modified by age and smoking status and were only reported for women [16], we conducted stratified analyses by age (<65 years, >=65 years), smoking status (current vs former smokers) and sex. There was an inverse trend for the Hannum measurements in men but not in women, where associations were null (Table 4). Associations for all three age acceleration measures were statistically significantly inversely associated with lung cancer in the younger (<65 years) but not older age group (Supplemental Table 1). Associations were similar among current and former smokers after adjusting for methylation predicted pack-years; associations were not estimated among never smokers due to small numbers (n=22 matched pairs; Supplemental Table 1). Finally, to examine whether associations might vary by histology, we separated NSCLC and SCLC; no associations were observed in either subgroup (data not shown).

**Table 4.** Odds ratios and 95% confidence intervals for the association between epigenetic age acceleration (using 3 different measures) and the risk of lung cancer in the CLUE II study, stratified by sex.

|  | Female |  | Male |  |
| --- | --- | --- | --- | --- |
|  | OR (95% CI) | p-value | OR (95% CI) | p-value |
| **AA_Hannum** |  |  |  |  |
| Q1 | 1 (ref.) |  | 1 (ref.) |  |
| Q2 | 0.74 (0.28, 2.00) | 0.56 | 0.78 (0.30, 2.00) | 0.60 |
| Q3 | 1.18 (0.49, 2.84) | 0.71 | 0.50 (0.21, 1.19) | 0.12 |
| Q4 | 0.79 (0.33, 1.88) | 0.60 | 0.29 (0.10, 0.80) | 0.017 |
|  |  | P trend = 0.78 |  | P trend = 0.015 |
| **AA_Horvath** |  |  |  |  |
| Q1 | 1 (ref.) |  | 1 (ref.) |  |
| Q2 | 0.92 (0.40, 2.13) | 0.84 | 0.72 (0.30, 1.73) | 0.47 |
| Q3 | 0.82 (0.34, 1.99) | 0.66 | 0.51 (0.21, 1.24) | 0.14 |
| Q4 | 0.92 (0.40, 2.15) | 0.85 | 0.50 (0.19, 1.35) | 0.17 |
|  |  | P trend = 0.80 |  | P trend = 0.13 |
| **AA_Pheno** |  |  |  |  |
| Q1 | 1 (ref.) |  | 1 (ref.) |  |
| Q2 | 0.70 (0.28, 1.76) | 0.44 | 1.11 (0.43, 2.86) | 0.83 |
| Q3 | 0.89 (0.38, 2.05) | 0.78 | 0.90 (0.36, 2.26) | 0.83 |
| Q4 | 0.93 (0.40, 2.16) | 0.86 | 0.60 (0.21, 1.74) | 0.35 |
|  |  | P trend = 0.97 |  | P trend = 0.30 |
| **IEAA_Hannum** |  |  |  |  |
| Q1 | 1 (ref.) |  | 1 (ref.) |  |
| Q2 | 1.27 (0.52, 3.08) | 0.60 | 0.31 (0.11, 0.82) | 0.018 |
| Q3 | 1.17 (0.47, 2.93) | 0.74 | 0.37 (0.14, 0.96) | 0.041 |
| Q4 | 0.92 (0.40, 0.10) | 0.84 | 0.35 (0.13, 0.92) | 0.034 |
|  |  | P trend = 0.68 |  | P trend = 0.03 |
| **IEAA_Horvath** |  |  |  |  |
| Q1 | 1 (ref.) |  | 1 (ref.) |  |
| Q2 | 0.85 (0.36, 2.00) | 0.70 | 0.85 (0.34, 2.15) | 0.73 |
| Q3 | 0.73 (0.31, 1.75) | 0.48 | 0.83 (0.33, 2.11) | 0.70 |
| Q4 | 0.21 (0.54, 2.74) | 0.64 | 0.53 (0.17, 1.58) | 0.25 |
|  |  | P trend = 0.65 |  | P trend = 0.28 |
| **IEAA_Pheno** |  |  |  |  |
| Q1 | 1 (ref.) |  | 1 (ref.) |  |
| Q2 | 1.01 (0.43, 2.36) | 0.98 | 1.34 (0.51, 3.52) | 0.55 |
| Q3 | 1.10 (0.46, 2.63) | 0.83 | 1.18 (0.46, 3.05) | 0.73 |
| Q4 | 1.04 (0.46, 2.35) | 0.92 | 0.63 (0.21, 1.82) | 0.39 |
|  |  | P trend = 0.88 |  | P trend = 0.38 |

IEAA: Intrinsic epigenetic age acceleration
\*Conditional logistic regression models, adjusting for batch effects, BMI, and smoking predicted packyears (using DNA methylation).

Prior studies suggest that epigenetic age acceleration may be strongly linked to the immune response, and specifically CD8 and CD4 naïve cells [40]. In this study, all three epigenetic clock measures were strongly associated with CD8 and CD4 naïve immune subsets in control subjects (Pearson correlations ranging from −0.22 to −0.41; Supplemental Table 2). The only statistically significant correlation between the clock measures and NK cells was for PhenoAge (r = −0.16); however, the NK cells in the reference library do not differentiate naïve and memory NK cells, so it is possible the associations would be different for NK naïve cells. The CD8 memory cells were positively associated with Hannum and Horvath clocks but not with PhenoAge. The IEAA measures (for each clock) were not associated with CD8 memory cells but were still strongly associated with CD8 naïve cells, which can be explain by the lack of adjustment for naïve and memory T cells as these fractions were not available in earlier deconvolution libraries (and memory cell represent a larger proportion of total T cells in older adults).

## Discussion

In this nested case-control study on incident lung cancer, we observed no positive associations between lung cancer risk and epigenetic age acceleration using three different measures (Horvath, Hannum and PhenoAge) with two adjustment approaches for each, i.e., intrinsic and extrinsic measures. We observed inverse associations for men and subjects below the median age, but not in women or older subjects.

Our null findings for epigenetic age acceleration associations with lung cancer risk using the Horvath and Hannum clocks are consistent with those reported in a nested case-control study in the Melbourne Collaborative Cohort Study (MCCS; 332 cases) [17]. Our null findings differ from those reported in the Women’s Health Initiative (WHI), where a 50% increase in risk of lung cancer was observed for every unit increase in intrinsic epigenetic age acceleration using the Horvath epigenetic age measure (p=3.4×10^−3^)[16]; it is worth noting that the number of lung cancer cases included in the WHI analysis was small (n=43). Our results for age acceleration based on the PhenoAge measure were also null, whereas a positive association was observed for PhenoAge and lung cancer risk in the MCCS (OR = 1.25, 95% CI = 1.05-1.49, for a 1 SD increase)[13]. The PhenoAge measure was derived using inflammatory phenotypes, in contrast to the other two epigenetic measures. Differences in the two populations may explain the different findings, such as smoking prevalence, although we could not confirm that as the MCCS analysis did not provide characteristics for the lung cancer case-control study (only for the pooled population). We observed that adjusting for methylation predicted pack-years attenuated the associations for PhenoAge in our analysis (among current smokers: before adjustment OR = 2.15, 95% CI = 0.92-5.04 for Q4 vs Q1; after adjustment OR = 1.40, 95% CI = 0.52-3.76; overall: before adjustment OR 1.22, 95% CI = 0.70-2.13; after adjustment OR = 0.86, 95% CI = 0.46-1.62). Thus, it is possible that elevated risk associated with the PhenoAge age acceleration in some studies is a measure of the residual effect of smoking, which is captured with the methylation markers for pack-years smoked.

The inverse associations we observed in men and subjects less than 65 years of age) were unexpected. It is possible these findings were due to chance, given that we performed several sub-analyses. Alternatively, these results may be due to a selection bias that occurred as a results of survivor bias in enrollment into our cohort (this could have occurrent if individuals with poor health and epigenetic age acceleration were less likely to participate).

Recent studies have begun to elucidate the biological processes that explain age acceleration associations detected using epigenetic clocks [40]. In a functional genomics study, changes in proportions of naive and activated immune blood cells were strongly associated with the Hannum and Horvath age acceleration measures [40]. We confirmed these relationships in our dataset using new immune cell reference libraries allowing the deconvolution of naive immune T cells [21]. The strong inverse correlations observed between naive T and B cells and the three age acceleration measurements suggest that they are strongly linked to changes in the immune response, which is not surprising, given that reduction of naïve T cells is a component of immunosenescence [41]. Of interest, the intrinsic epigenetic age acceleration (IEAA) measurements remained strongly associated with the CD8 naïve cells; future analyses using intrinsic measures of age acceleration should adjust for these cells.

The strengths of our study include the prospective nature of the analysis with a long follow-up period, thus removing the potential for spurious associations that may be driven by the cancer progression (i.e., reverse causation), a relatively large sample size for methylation analyses, and tight adjustment for smoking. In addition, the cancer ascertainment for the CLUE II cohort is very high, given the quality of cancer registry data. The limitations of our analysis include one-time point for epigenetic measurements and lack of data on non-Whites, thus limiting generalizability.

To our knowledge, this is the third prospective study examining the association between epigenetic aging, measured in peripheral blood, and risk of lung cancer. Findings from this study suggest that there are no strong positive associations between biological aging, measured an average of 15 years prior to cancer, and lung cancer risk. The majority of cases in this study were ever smokers (90%), and smoking history was well controlled for, suggesting that biological aging, independent of smoking, is not associated with an increased risk of lung cancer, at least among smokers. Our data also suggest that prior associations with PhenoAge and lung cancer might have been due to residual effects of smoking. Future studies should include more racially diverse populations and examine associations among never smokers.

## Data Availability

The datasets generated during the current study are available from the corresponding author on reasonable request.

## Acknowledgements

Cancer data were provided by the Maryland Cancer Registry, Center for Cancer Prevention and Control, Maryland Department of Health, with funding from the State of Maryland and the Maryland Cigarette Restitution Fund. The collection and availability of cancer registry data is also supported by the Cooperative Agreement NU58DP006333, funded by the Centers for Disease Control and Prevention. Its contents are solely the responsibility of the authors and do not necessarily represent the official views of the Centers for Disease Control and Prevention or the Department of Health and Human Services.

---

**Supplemental Table 1.** Odds ratios and 95% confidence intervals for the association between epigenetic age acceleration (using 3 different measures) and the risk of lung cancer in the CLUE II study, stratified by age and smoking status.

|  | OR (95% CI)* | p-value | OR (95% CI)* | p-value |
| --- | --- | --- | --- | --- |
|  | Age <65 years |  | Age ≥ 65 years |  |
| AA_Hannum | 0.63 (0.44, 0.90) | 0.011 | 0.90 (0.53, 1.52) | 0.69 |
| AA_Horvath | 0.62 (0.44, 0.88) | 0.007 | 1.10 (0.69, 1.75) | 0.70 |
| AA_Pheno | 0.76 (0.58, 1.00) | 0.046 | 1.07 (0.75, 1.52) | 0.72 |
| IEAA_Hannum | 0.69 (0.48, 1.00) | 0.05 | 0.91 (0.52, 1.59) | 0.74 |
| IEAA_Horvath | 0.73 (0.52, 1.04) | 0.08 | 1.08 (0.67, 1.74) | 0.74 |
| IEAA_Pheno | 0.74 (0.55, 0.98) | 0.035 | 1.13 (0.77, 1.67) | 0.52 |
|  | Current smoker** |  | Former smoker** |  |
| AA_Hannum | 0.76 (0.49, 1.20) | 0.24 | 0.69 (0.44, 1.07) | 0.09 |
| AA_Horvath | 0.68 (0.45, 1.02) | 0.06 | 0.82 (0.54, 1.27) | 0.38 |
| AA_Pheno | 0.93 (0.68, 1.28) | 0.66 | 0.84 (0.61, 1.16) | 0.30 |
| IEAA_Hannum | 0.93 (0.58, 1.48) | 0.76 | 0.71 (0.45, 1.12) | 0.14 |
| IEAA_Horvath | 0.81 (0.53, 1.22) | 0.31 | 0.91 (0.58, 1.43) | 0.69 |
| IEAA_Pheno | 1.02 (0.73, 1.44) | 0.90 | 0.85 (0.60, 1.20) | 0.36 |

IEAA: Intrinsic epigenetic age acceleration
\*Conditional logistic regression models, adjusting for batch effects, BMI, and smoking predicted packyears (using DNA methylation).
\*\*Never smokers were not included in this analysis as there were too few.

**Supplemental Table 2.** Correlations between epigenetic clocks and selected immune cell proportions (estimated from methylation data). Correlations for CD4 memory and B cell memory are not shown as the correlations were statistically significant.

|  | **AA**<br>**Hannum** | **IEAA**<br>**Hannum** | **AA**<br>**Horvath** | **IEAA**<br>**Horvath** | **AA**<br>**PhenoAge** | **IEAA**<br>**PhenoAge** |
| --- | --- | --- | --- | --- | --- | --- |
| **CD4 naïve** | -0.3677 | -0.1935 | -0.1884 | -0.0011 | -0.2249 | -0.1104 |
| **p values** | <0.0001 | 0.0051 | 0.0066 | 0.9877 | 0.0011 | 0.1124 |
| **CD8 naïve** | -0.4245 | -0.4076 | -0.2305 | -0.1706 | -0.32 | -0.3196 |
| **p values** | <0.0001 | <0.0001 | 0.0008 | 0.014 | <0.0001 | <0.0001 |
| **CD8 memory** | 0.3316 | 0.1264 | 0.3202 | -0.0026 | -0.0203 | 0.098 |
| **p values** | <0.0001 | 0.0689 | <0.0001 | 0.9706 | 0.7713 | 0.1591 |
| **B cell naïve** | -0.2507 | -0.1485 | -0.1786 | -0.0609 | -0.3272 | -0.1856 |
| **p values** | 0.0003 | 0.0323 | 0.01 | 0.3831 | <0.0001 | 0.0073 |

IEAA: Intrinsic epigenetic age acceleration

## References

1. Sung H, Ferlay J, Siegel RL, Laversanne M, Soerjomataram I, Jemal A, Bray F: Global Cancer Statistics 2020: GLOBOCAN Estimates of Incidence and Mortality Worldwide for 36 Cancers in 185 Countries. CA: a cancer journal for clinicians 2021, 71(3):209–249.

2. Bosse Y, Amos CI: A Decade of GWAS Results in Lung Cancer. Cancer epidemiology, biomarkers & prevention : a publication of the American Association for Cancer Research, cosponsored by the American Society of Preventive Oncology 2018, 27(4):363–379.

3. Lebrett MB, Crosbie EJ, Smith MJ, Woodward ER, Evans DG, Crosbie PAJ: Targeting lung cancer screening to individuals at greatest risk: the role of genetic factors. J Med Genet 2021, 58(4):217–226.

4. Colson YL, Shepard JO, Lennes IT: New USPSTF Guidelines for Lung Cancer Screening: Better but Not Enough. JAMA Surg 2021, 156(6):513–514.

5. Michaud DS, Kelsey KT: DNA Methylation in Peripheral Blood: Providing Novel Biomarkers of Exposure and Immunity to Examine Cancer Risk. Cancer epidemiology, biomarkers & prevention : a publication of the American Association for Cancer Research, cosponsored by the American Society of Preventive Oncology 2021, 30(12):2176–2178.

6. Baglietto L, Ponzi E, Haycock P, Hodge A, Bianca Assumma M, Jung CH, Chung J, Fasanelli F, Guida F, Campanella G et al: DNA methylation changes measured in pre-diagnostic peripheral blood samples are associated with smoking and lung cancer risk. International journal of cancer Journal international du cancer 2017, 140(1):50–61.

7. Heikkinen A, Bollepalli S, Ollikainen M: The potential of DNA methylation as a biomarker for obesity and smoking. J Intern Med 2022.

8. Horvath S, Zhang Y, Langfelder P, Kahn RS, Boks MP, van Eijk K, van den Berg LH, Ophoff RA: Aging effects on DNA methylation modules in human brain and blood tissue. Genome biology 2012, 13(10):R97.

9. Marioni RE, Shah S, McRae AF, Chen BH, Colicino E, Harris SE, Gibson J, Henders AK, Redmond P, Cox SR et al: DNA methylation age of blood predicts all-cause mortality in later life. Genome biology 2015, 16:25.

10. Chen BH, Marioni RE, Colicino E, Peters MJ, Ward-Caviness CK, Tsai PC, Roetker NS, Just AC, Demerath EW, Guan W et al: DNA methylation-based measures of biological age: meta-analysis predicting time to death. Aging 2016, 8(9):1844–1865.

11. Roetker NS, Pankow JS, Bressler J, Morrison AC, Boerwinkle E: Prospective Study of Epigenetic Age Acceleration and Incidence of Cardiovascular Disease Outcomes in the ARIC Study (Atherosclerosis Risk in Communities). Circ Genom Precis Med 2018, 11(3):e001937.

12. Perna L, Zhang Y, Mons U, Holleczek B, Saum KU, Brenner H: Epigenetic age acceleration predicts cancer, cardiovascular, and all-cause mortality in a German case cohort. Clinical epigenetics 2016, 8:64.

13. Dugue PA, Bassett JK, Wong EM, Joo JE, Li S, Yu C, Schmidt DF, Makalic E, Doo NW, Buchanan DD et al: Biological Aging Measures Based on Blood DNA Methylation and Risk of Cancer: A Prospective Study. JNCI Cancer Spectr 2021, 5(1).

14. Huang RC, Lillycrop KA, Beilin LJ, Godfrey KM, Anderson D, Mori TA, Rauschert S, Craig JM, Oddy WH, Ayonrinde OT et al: Epigenetic Age Acceleration in Adolescence Associates With BMI, Inflammation, and Risk Score for Middle Age Cardiovascular Disease. J Clin Endocrinol Metab 2019, 104(7):3012–3024.

15. Nannini DR, Joyce BT, Zheng Y, Gao T, Liu L, Yoon G, Huan T, Ma J, Jacobs DR, Jr., Wilkins JT et al: Epigenetic age acceleration and metabolic syndrome in the coronary artery risk development in young adults study. Clinical epigenetics 2019, 11(1):160.

16. Levine ME, Hosgood HD, Chen B, Absher D, Assimes T, Horvath S: DNA methylation age of blood predicts future onset of lung cancer in the women’s health initiative. Aging 2015, 7(9):690–700.

17. Dugue PA, Bassett JK, Joo JE, Jung CH, Ming Wong E, Moreno-Betancur M, Schmidt D, Makalic E, Li S, Severi G et al: DNA methylation-based biological aging and cancer risk and survival: Pooled analysis of seven prospective studies. International journal of cancer Journal international du cancer 2018, 142(8):1611–1619.

18. Comstock GW, Helzlsouer KJ, Bush TL: Prediagnostic serum levels of carotenoids and vitamin E as related to subsequent cancer in Washington County, Maryland. The American journal of clinical nutrition 1991, 53(1 Suppl):260S–264S.

19. Braun MM, Helzlsouer KJ, Hollis BW, Comstock GW: Colon cancer and serum vitamin D metabolite levels 10-17 years prior to diagnosis. American journal of epidemiology 1995, 142(6):608–611.

20. Zhao N, Ruan M, Koestler DC, Lu J, Salas LA, Kelsey KT, Platz EA, Michaud DS: Methylation-derived inflammatory measures and lung cancer risk and survival. Clinical epigenetics 2021, 13(1):222.

21. Salas LA, Zhang Z, Koestler DC, Butler RA, Hansen HM, Molinaro AM, Wiencke JK, Kelsey KT, Christensen BC: Enhanced cell deconvolution of peripheral blood using DNA methylation for high-resolution immune profiling. Nature communications 2022, 13(1):761.

22. Koestler DC, Jones MJ, Usset J, Christensen BC, Butler RA, Kobor MS, Wiencke JK, Kelsey KT: Improving cell mixture deconvolution by identifying optimal DNA methylation libraries (IDOL). BMC bioinformatics 2016, 17(1):120.

23. Salas LA, Koestler DC, Butler RA, Hansen HM, Wiencke JK, Kelsey KT, Christensen BC: An optimized library for reference-based deconvolution of whole-blood biospecimens assayed using the Illumina HumanMethylationEPIC BeadArray. Genome biology 2018, 19(1):1–14.

24. Aryee MJ, Jaffe AE, Corrada-Bravo H, Ladd-Acosta C, Feinberg AP, Hansen KD, Irizarry RA: Minfi: a flexible and comprehensive Bioconductor package for the analysis of Infinium DNA methylation microarrays. Bioinformatics 2014, 30(10):1363–1369.

25. Fortin JP, Triche TJ, Jr., Hansen KD: Preprocessing, normalization and integration of the Illumina HumanMethylationEPIC array with minfi. Bioinformatics 2017, 33(4):558–560.

26. Triche TJ, Jr., Weisenberger DJ, Van Den Berg D, Laird PW, Siegmund KD: Low-level processing of Illumina Infinium DNA Methylation BeadArrays. Nucleic Acids Res 2013, 41(7):e90.

27. Xu Z, Niu L, Li L, Taylor JA: ENmix: a novel background correction method for Illumina HumanMethylation450 BeadChip. Nucleic Acids Res 2016, 44(3):e20.

28. Teschendorff AE, Marabita F, Lechner M, Bartlett T, Tegner J, Gomez-Cabrero D, Beck S: A beta-mixture quantile normalization method for correcting probe design bias in Illumina Infinium 450 k DNA methylation data. Bioinformatics 2013, 29(2):189–196.

29. Pidsley R, Cc YW, Volta M, Lunnon K, Mill J, Schalkwyk LC: A data-driven approach to preprocessing Illumina 450K methylation array data. BMC genomics 2013, 14:293.

30. Teschendorff AE, Zhuang J, Widschwendter M: Independent surrogate variable analysis to deconvolve confounding factors in large-scale microarray profiling studies. Bioinformatics 2011, 27(11):1496–1505.

31. Leek JT, Johnson WE, Parker HS, Jaffe AE, Storey JD: The sva package for removing batch effects and other unwanted variation in high-throughput experiments. Bioinformatics 2012, 28(6):882–883.

32. Leek JT, Storey JD: Capturing heterogeneity in gene expression studies by surrogate variable analysis. PLoS genetics 2007, 3(9):1724–1735.

33. Sugden K, Hannon EJ, Arseneault L, Belsky DW, Broadbent JM, Corcoran DL, Hancox RJ, Houts RM, Moffitt TE, Poulton R et al: Establishing a generalized polyepigenetic biomarker for tobacco smoking. Transl Psychiatry 2019, 9(1):92.

34. Hannum G, Guinney J, Zhao L, Zhang L, Hughes G, Sadda S, Klotzle B, Bibikova M, Fan JB, Gao Y et al: Genome-wide methylation profiles reveal quantitative views of human aging rates. Mol Cell 2013, 49(2):359–367.

35. Horvath S: DNA methylation age of human tissues and cell types. Genome biology 2013, 14(10):R115.

36. Levine ME, Lu AT, Quach A, Chen BH, Assimes TL, Bandinelli S, Hou L, Baccarelli AA, Stewart JD, Li Y et al: An epigenetic biomarker of aging for lifespan and healthspan. Aging (Albany NY) 2018, 10(4):573–591.

37. Houseman EA, Accomando WP, Koestler DC, Christensen BC, Marsit CJ, Nelson HH, Wiencke JK, Kelsey KT: DNA methylation arrays as surrogate measures of cell mixture distribution. BMC bioinformatics 2012, 13:86.

38. Salas LA, Koestler DC, Butler RA, Hansen HM, Wiencke JK, Kelsey KT, Christensen BC: An optimized library for reference-based deconvolution of whole-blood biospecimens assayed using the Illumina HumanMethylationEPIC BeadArray. Genome biology 2018, 19(1):64.

39. Sugden K, Hannon EJ, Arseneault L, Belsky DW, Broadbent JM, Corcoran DL, Hancox RJ, Houts RM, Moffitt TE, Poulton R: Establishing a generalized polyepigenetic biomarker for tobacco smoking. Translational psychiatry 2019, 9(1):1–12.

40. Jonkman TH, Dekkers KF, Slieker RC, Grant CD, Ikram MA, van Greevenbroek MMJ, Franke L, Veldink JH, Boomsma DI, Slagboom PE et al: Functional genomics analysis identifies T and NK cell activation as a driver of epigenetic clock progression. Genome biology 2022, 23(1):24.

41. Santoro A, Bientinesi E, Monti D: Immunosenescence and inflammaging in the aging process: age-related diseases or longevity? Ageing Res Rev 2021, 71:101422.

